# Examining the Relationship between COVID-19 Vaccinations and Reported Incidence

**DOI:** 10.1101/2021.06.30.21259794

**Authors:** Enbal Shacham, Stephen Scroggins, Alexander Garza

## Abstract

As COVID-19 has caused significant morbidity and mortality throughout the world, the development and distribution of an effective vaccine have been swift but not without challenges. Earlier demand and access barriers have seemingly been addressed with more free and accessible vaccines now available for a wide variety of ages. While rates of COVID-19 have decreased overall, some geographic areas continue to experience rapid outbreaks. The purpose of this study was to examine the relationship between vaccination uptake and weekly COVID-19 cases throughout locations in the state of Missouri.

**Methods:** Among all Missouri counties and two cities (n=117), weekly COVID-19 incidence and cumulative proportion of residents fully vaccinated were abstracted from the Missouri Department of Health and Senior Services during a 25-week period from January 4 to Jun 26, 2021. Additional ecological variables known to be associated with COVID-19 incidence and prevalence were collected from the U.S. Census Bureau and integrated into data: total population, proportion of nonwhite residents, annual median household income, proportion of residents working in public facing occupations. Descriptive and inferential statistics were completed which included the calculation of both linear and nonlinear models using repeated measure data to determine the quantitative association between vaccination uptake and reported COVID-19 cases in the presence of location characteristics.

**Results:** Throughout the 25 weeks of observations, the average weekly number of COVID-19 cases reported was 66.1 (SD=260.8) while the average cumulative proportion vaccinated individuals at the end of the 25 weeks was 25.8% (SD=6.8%) among study locations. While graphing seemed to suggest a more nonlinear relationship between COVID-19 incidence and proportion vaccinated, comparison of crude linear and nonlinear models pointed to the relationship likely being linear during study period. The final adjusted linear model exhibited a significant relationship between COVID-19 cases and proportion vaccinated, specifically every percent increase in population vaccinated resulted in 3 less weekly COVID-19 cases being reported (β -3.74, p<0.001. Additionally, when controlling for other factors, the adjusted model revealed locations with higher proportions of nonwhite residents were likely to experience less weekly COVID-19 cases (β -1.48, p=0.037).

**Discussion:** Overall, this study determined that increasing the proportion of residents vaccinated decreases COIVD-19 cases by a substantial amount over time. These findings provide insights into possible messaging strategies that can be leveraged to develop more effective implementation and uptake. As the COVID-19 pandemic persists and vaccination numbers begin to plateau, diverse communication strategies become a critical necessity to reach a wider population.

## Introduction

The COVID-19 pandemic has caused significant morbidity and mortality throughout the world. With continued new daily infections and the roll-out of vaccinations, there are dropping infections in geographic regions where there are higher rates of vaccination.^1^ As expected, vaccinations are working to reduce new infections as they provide protection for individuals who are able to access and receive them.^2^ The concerns that COVID-19 variants limit the efficacy and effectiveness of COVID-19 vaccines have been expressed and these fears have yet to be substantiated; as additional research needs to continue to monitor rates of infection, COVID-19 variants, and rates of vaccinations for each community.

Herd immunity, the effort to achieve COVID-19 immunity has been discussed extensively in the literature and media as an effort to overcome COVID-19.^3,4^ This type of immunity could occur with a combination of individuals who have been infected with COVID-19 and those who have been vaccinated. It continues to be unclear how long either immunity persists.^3^ Yet, even more importantly, the size or geography of the “herd” has been very difficult to measure in the U.S. with many different COVID-19 mitigation strategies being implemented in different geographies: cities, counties, and states, with no limitations of travel between them. Thus, the measure of what potential herd immunity may occur is unclear in the U.S., while other countries may be achieving this measure of COVID-19 management. ^3,5^

Furthermore, there are continued challenges with the uptake of the COVID-19 vaccine in the U.S. While demand and access seem to have been solved in many ways, there are now more free and accessible vaccines for all ages who have been FDA-emergency authorized approved. Many of these barriers to vaccination include little effective communicated benefits and safety, and overall mistrust of the public health and healthcare system that have been documented with non-white communities as well as white, rural residents.^6^ These reasons for non-vaccination differ by community, and as such, promotion efforts should respond appropriately to each barrier. Lastly, there continues to be a need to increase ease of vaccine access, which may include workplace and community-based vaccinations, such as churches, professional sports events, and paid time leave to get vaccinated that may provide greater incentive with very little access challenges.

Increasing vaccination rates by community and population is imperative to achieve more immunity in our communities. In the U.S., there have been documented reduction in COVID-19 infection rates, yet this is not an equal measure across all populations.^7^ Missouri provides a diverse population as it experiences the barriers to uptake in more urban communities among Black and African American residents as well as white and Republican voters in the more rural areas.^6,8-10^ Thus, examining cases of infection and vaccination across these regions may guide how herd immunity may occur across regions like Missouri. The purpose of this study was to assess the relationship between COVID-19 cases and the proportion of vaccination in Missouri.

## Methods

This study utilized an ecological framework at the county-level as the base observation, except in two instances where a city was separated out from their respective county. Using this level of observation, the final sample consisted of 115 Missouri counties and two Missouri cities for a final sample of 117. Weekly observations among the sample began on January 4, 2021 until June 26, 2021 for a temporal period of 25 weeks. The study period reflects the availability of the COVID-19 vaccine and current dates^11^.

Both the primary independent variable and the study outcome were obtained from the Missouri Department of Health and Senior Services’ (MDHSS) public COVID-19 dashboard application and data repository^12^. The study outcome was defined as weekly number of new COVID-19 cases across locations. Data were aggregated weekly, rather than daily, to limit bias in respective county reporting protocols.

The primary independent variable for this study was the cumulated weekly percent estimated of residents fully vaccinated. Cumulated proportion, rather than number or proportion of newly vaccinated, was used due to the accumulated protection offered by vaccines during the study period. Covariates, hypothesized to also have an influence on weekly number of new COVID-19 cases reported, were matched to location, and integrated into data. These covariates included: total population of residents, proportion of nonwhite residents, annual median household income, proportion of the population working in food service and proportion of those working in healthcare support as defined by the U.S. Department of Labor^13^. These two occupation categories were chosen based on required public interaction and previously identified increased transmission risk^14^.

First, descriptive statistics were completed primarily to determine significant bivariate associations between COVID-19 cases and location characteristics. To accomplish this, a cross-sectional snapshot was taken; COVID-19 cases were averaged across weeks among each location (n=117). Using this data structure was determined to be appropriate given the time-independent and stationary location characteristic variables across the study period.

Significant associations between averaged COVID-19 cases and total population, proportion of non-white residents, proportion healthcare support workers, proportion food-service workers, median annual income, and final week cumulative proportion vaccinated residents were determined, respectively, between locations using Spearman rank correlation.

Next, COVID-19 cases and cumulative proportion of residents vaccinated were averaged across all locations by each week (n=25) and plotted to better identify overall trends between COVID-19 cases and proportion vaccinated. Both a linear and non-linear trendlines were applied and used to deduce overall fit.

Based on preliminary findings from correlations and plotting, a repeated measure data structure using weekly COVID-19 cases among each location (n=2,925) was used to fit both a linear and non-linear regression curves in order to determine a more accurate relationship between weekly COVID-19 cases and cumulative proportion of residents vaccinated. Due to the serial correlation present within cumulated vaccinated variable, a general estimating equation framework with an autoregressive working correlation matrix was used for all regression models. For the nonlinear model, a log-level or exponential regression model was used and then back-transformed for interpretability.

The crude linear and non-linear model were then assessed using quasi-Akaike’s information criterion (QIC) developed by Pan^15^. Based on compared QIC an adjusted model was then developed using the best fit (linear vs. nonlinear) regression to describe the relationship more realistically between weekly COVID-19 cases and cumulative population vaccinated in the presence of the above aforementioned covariates.

All statistics were completed in R software for statistical computing and all significance was determined at alpha = 0.05^16^. All ethical guidelines were followed according to the Helsinki Declaration and design and protocols was determined exempt by Institutional Review Board.

## Results

Throughout the 25 weeks of observations, the average weekly new COVID-19 cases reported by locations was 66.1 (SD=260.8). The median number of new cases reported weekly by county was calculated to be 11 with a minimum of 0 and a maximum of 6336 cases reported. The average cumulative vaccinated population at the end of the 25 weeks was 25.8% (SD=6.8%) per county with a median of 25.4% and a minimum and maximum of 11.9% and 45.5%, respectively, among Missouri locations (n=117).

Significant correlations were identified between average weekly COVID-19 cases and all characteristics previously determined to be associated with COVID-19 prevalence (Table 1). Specifically, locations with higher populations (p<0.001), higher proportion of non-white residents (p<0.001), higher proportion of food service-workers (p<0.001), and higher median annual income (p<0.001) were significantly more likely to report higher average weekly COVID-19 cases. Conversely, locations with higher proportions of healthcare support workers (p<0.001) and higher proportion of vaccinated residents (p<0.001) were more likely to report significantly less average weekly COVID-19 cases during the 25-week period.

**Table 1.** Bivariate correlation among location characteristics and weekly average COVID-19 cases among locations across the state of Missouri from January 4 to June 26, 2021 (n=117)

| | Average weekly COVID-19 cases ($\rho$)<sup>a</sup> |
| --- | --- |
| Total population | 0.66*** |
| Proportion nonwhite residents | 0.42*** |
| Proportion healthcare support worker | -0.24*** |
| Proportion food service worker | 0.20*** |
| Median annual household income | 0.29*** |
| Cumulative proportion vaccinated | -0.20*** |

Significance indicated at 0.05\*, 0.01\*\*, 0.001\*\*\*
a. $\rho$ coefficient calculated using Spearman rank correlation test

The average reported number of COVID-19 cases and the average cumulative proportion of vaccinated residents across all study locations for each week were calculated and plotted in Figure 1. This figure suggests an inverse and possibly nonlinear relationship exists between COVID-19 cases and proportion vaccinated. According to this aggregated data, a nonlinear trendline appear to fit somewhat better. This occurrence was tested along with more specific relationships determined using regression models in Table 2.

**Table 2.** Crude and adjusted linear and log-linear regression models predicting number of weekly new COVID-19 cases among Missouri locations (n=117) over a 25-week period.

|  | Linear Model 1<sup>a</sup> | Log-Level Model 2<sup>b</sup> | Linear Model 3<sup>c</sup> |
| --- | --- | --- | --- |
| | Crude $\beta$ | Crude $\beta$ | Adjusted $\beta$ |
| Intercept | **111.53\*\*\*** | **3.02\*\*\*** | 0.83 |
| Cumulative proportion of vaccinated population | **-2.74\*\*\*** | **-0.04\*\*\*** | **-3.74\*\*\*** |
| Total population |  |  | **0.01\*\*\*** |
| Proportion nonwhite residents |  |  | **-1.48\*** |
| Proportion healthcare support workers |  |  | 1.82 |
| Proportion food service workers |  |  | 2.62 |
| Median annual household income |  |  | 0.01 |

a. Quasi-loglikelihood = -1403, Quasi-Akaike's information criterion = 2825.8
b. Quasi-loglikelihood = -1403, Quasi-Akaike's information criterion = 2829.2
c. Quasi-loglikelihood = -1400, Quasi-Akaike's information criterion = 2825.3

**Figure 1.**
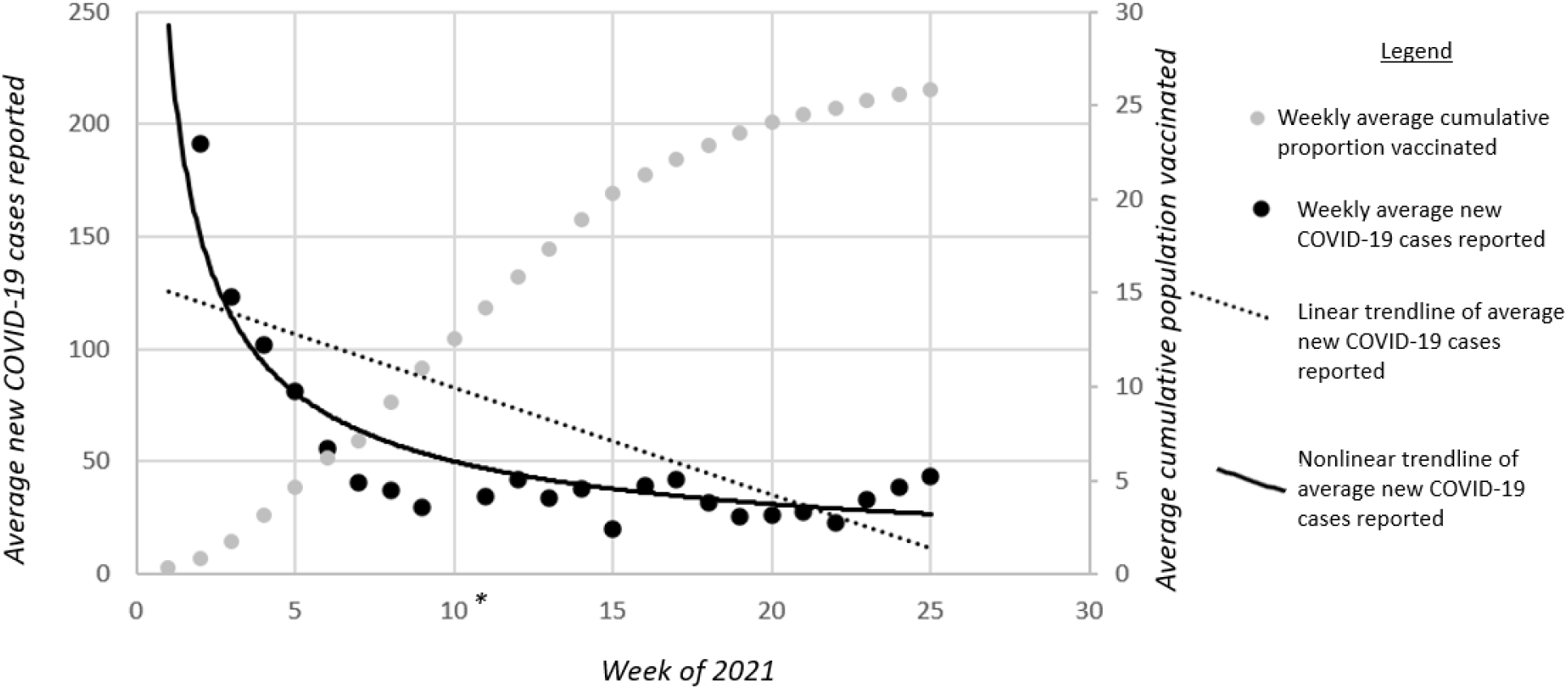
Weekly average of cumulative proportion vaccinated residents and average COVID-19 cases reported among Missouri locations across 25 weeks *Average Week 10 COVID-19 cases not shown at 428 cases.

Model 1 of Table 2 reveals a crude linear regression that proposes for every percent increase in vaccination uptake among locations, there is like to be 3 less COVID-19 cases reported (β - 2.74, p<0.001). In contrast Model 2, which uses a log-level, or exponential approach reveals that for every percent increase in population vaccinated among locations, COVID-19 cases will decrease by 4% (β 0.04, p<0.001). While these two crude models suggest very similar outcomes, comparing QIC’s suggest the linear model is, if only very slightly, a better fit with a QIC of 2825.8 compared the nonlinear model QIC of 2829.3. Thus, the final adjusted model used a linear framework to predict COVID-19 cases from proportion vaccinated controlling for covariates.

When controlling for social and geo-characteristics, every percent increase in vaccinated population is likely to result in nearly 4 less reported COVID-19 cases across locations (β -3.74, p<0.001). Additionally, this model suggest, unsurprisingly, that locations with larger populations are likely to report higher number of COVID-19 cases (β 0.01, p<0.001). Surprisingly however, when controlling for other factors, location with higher proportions of non-white residents were likely to experience significantly less weekly COVID-19 cases (β -1.48, p<0.032). This is contrasted with the aggregated tests featured in Table 1 which suggest without adjusting for covariates locations with higher proportions of non-white residents were likely to experience a higher average number of COVID-19 cases over the course of the study period.

## Discussion

The purpose of this study was to examine the relationship between COVID-19 infection and vaccination rates within a diverse state that has expressed vaccine and mask-wearing hesitancy, and paradoxically supported COVID-19 mitigation strategies and vaccine seeking. This study identified that the increase in vaccination reduced the infection rate in a linear manner. Thus, suggesting that each person getting vaccinated has an impact in supports their individual and community health. The predictors of infection persisted, as they had throughout the pandemic with higher population density and areas where more front-line workers live.

Focus on increased vaccine uptake in more populated communities is urgent. Additionally, understanding that rural and urban communities are connected geographically in states like Missouri is important to consider in the response to community health needs. These findings identify that vaccination and COVID-19 rates are conversely associated with county-level sociodemographic characteristics. These results can help identify further purposeful and focused intervention on job sites and continued in more urban environments. These efforts have been delayed, much of the medical and vaccine mistrust has been blamed on the long history of use of Black and Brown community members in the U.S.,^10,17^ yet the continued burden of COVID-19 that has been experienced in Black and Brown residents throughout the country persist and little changes have been implemented to reduce the impact of COVID-19 on the exposure risk and health care needs of communities of color. Thus, specific efforts to overcome the systemic mistrust are necessary, for many reasons, and particularly to increase the rates of vaccination as a method to protect themselves and community members.

Further, the distrust that rural residents and Republicans have expressed demand a different type of intervention and communication strategy. While data have been promoted much of the social media-driven communication that had driven the distrust and anger about COVID-19 mitigation strategies are documented to be rooted in few people yet have proliferated widely.^18^ While important to document to better understand how to prevent future challenges, these reports may not assist in changing beliefs and practices. Better efforts to support how social networks may serve to influence vaccine uptake is likely beneficial in both rural and urban communities.

As the U.S. has not implemented federal mandates for COVID-19 mitigation or vaccination strategies, thus, leaving states to respond either proactively or reactively to COVID-19. The diverse, and at times divisive, responses by residents in states that experience higher divergence in their risk perceptions have been challenged to have a comprehensive and community-wide response. Promoting the health of a community has not previously felt so divisive, yet perhaps it has always been as the founding of this country, we just had not experienced the efficiency in inequities as swiftly as COVID-19 has them pronounced.

This study highlights that herd immunity is not achievable within an acceptable time frame that does not cause significant morbidity and mortality. This study period spanned approximately 6 months over times of limited access to and more accessible vaccinations. During this study period, additional segments of the population had also been approved to be vaccinated. These results highlight the opportunity to craft public health messaging to encourage vaccination by using language that highlights each person who is fully vaccinated reduces the likelihood of about 2 COVID-19 infections. During a time where there is significant hesitation for required vaccination, this message may provide great support for public health and health care practitioners.

## Data Availability

All raw data used for this study is available online through the Missouri Department of Health and Human Services and through the U.S. Census Bureau.

## Acknowledgements

This study was made possible with support from Saint Louis University College for Public Health and Social Justice, The Geospatial Institute, and The Sinquefield Center for Applied Economic Research.

